# Gender influence on changes of Coagulation factors in cryoprecipitate plasma at Kisii Teaching and Referral Hospital

**DOI:** 10.1101/2023.05.05.23289552

**Authors:** Collince Odiwuor Ogolla, Benson Nyanchongi, Chrisphine Momanyi

## Abstract

**Background:** Cryoprecipitate is used without knowing the concentration of coagulation factors it contains which poses danger of circulatory overload with no any improvement if several bags with low factor levels are transfused when the levels changes or drastically reduce.

**Objective:** The objective was to evaluate gender influence on changes of Coagulation factors in cryoprecipitate plasma at Kisii Teaching and Referral Hospital.

**Methods:** The study involved time series analysis design involving analysis of cryoprecipitate during storage at -18°C for 5 weeks. Blood collected from the Kisii satellite blood transfusion center was received at Hematology laboratory where factor assays were performed on Erba Mannheim ECL 105 semi-automated coagulation analyzer. Thawing for subsequent coagulation factor analysis and serial testing was done using Stericox Plasma Thawing Bath at 37°C, for 45 mins. Data were entered into Excel and analyzed by SPSS version 25.

**Results:** The mean rank for cryoprecipitate from week one to week five was in a decreasing trend with 3.00, 1.99 and 1.01 respectively. This confirms a steady statistical significant difference in mean ranks for the time period. The coagulation factors in cryoprecipitate plasma in the donated blood were not affected by the gender of the blood donor.

**Conclusion:** The gender of the donor has no effect on the coagulation factors in cryoprecipitate plasma

## Introduction

Cryoprecipitate is a frozen blood product prepared by thawing of FFP to 1°C to 6 °C and thereafter centrifuged, the precipitate is collected which is the cryoprecipitate (2). Cryo is rich in clotting factors which are proteins which help to slow or stop bleeding hence reducing blood loss. The factors include FI, FXIII, FVIII and vWF. Cryoprecipitate is used for the control and stoppage of bleeding in people in cases where their blood doesn’t properly clot. This comprises patients experiencing grave but rare inherited disorders like Haemophilia A (deficiency of FVIII) and also von Willebrand disease (deficiency of vWF) (1). Cryoprecipitate has high concentration of coagulation FVIII, FXIII, and FI (2).

Cryoprecipitate has similar storage condition as fresh frozen plasma (−18°C for 1 year) but cannot be re-frozen once separated as FFP (3). Once thawed, it is stored at room temperature for 4 hrs. Cryoprecipitate is indicated for treatment for FVIII deficiency, congenital or acquired FI deficiency, treatment for von Willebrand’s Disease, FXIII deficiency, “Fibrin Glue” which is smeared to surgical sites (3). Multiple reports have demonstrated the associations of VWF and FVIII with race, gender, and age. Mean levels of VWF and FVIII are significantly higher in females than in males and in African-American than in Caucasians (4)

There are several resources-limited county hospitals, referral hospitals and sub-county health centers laboratories in Kenya where factor assay are not performed systematically on patients experiencing bleeding disorders attending these facilities, cryoprecipitate and FFP is used without knowing the concentration of coagulation factors it contain which might cause circulatory overload with no any improvement if several bags are transfused with low level of coagulation factors (5). Blood components transfusion especially cryoprecipitate and FFP is also carried out without prior evaluating the levels, content or quality of coagulation factors which poses risk in efficient patient management at Kisii Teaching and Referral Hospital. Changes occurring in coagulation factors in stored cryoprecipitate have not been analyzed to evaluate its association with gender and thus, this study aimed at evaluating gender influence on changes of Coagulation factors in cryoprecipitate plasma at Kisii Teaching and Referral Hospital.

## Methods

### Study Site

This study was conducted at Kisii Teaching and referral hospital (KTRH) laboratory department. KTRH is located within Kisii town at the southern end of the western Kenyan highlands at an altitude of 1,660m above sea level. Coordinates for the town are 0°41’S 34°46’E / 0.683°S 34.767°E

### Sample Size

The study involved 108 eligible volunteer blood donors at Kisii Satellite Blood Transfusion Center, who met the donor suitability criteria following the World Health Organization guidelines.

### Study Design

This study involved time series analysis design involving time series analysis of cryoprecipitate plasma during storage at -18°C for 5 weeks at an interval of one week. Four hundred- and fifty- ml blood was collected into tetra blood bags containing citrate-phosphate-adenine anticoagulant- preservative (*CPDA*-*1*) as an anti-coagulant preservative for subsequent processing into cryoprecipitate for storage at -18°C. The collected blood was centrifuged at 4000 RPM for 9 minutes within 5 - 8 hours after collection in a separate sanitized room where about 180ml plasma was formed as supernatant which then was separated and collected. The 180ml plasma obtained through centrifugation was aliquoted in three parts each containing 60ml. The first aliquot was used to assess the changes in coagulation factors in cryoprecipitate plasma at room temp at baseline during week one of collection (baseline), the second aliquot was used to assess the changes in coagulation factors in cryoprecipitate plasma storage at -18°C temp after three weeks of storage, the third aliquot was used to assess the changes in coagulation factors in cryoprecipitate plasma storage at -18°C temp after five weeks of storage. Coagulation factor analysis was performed using Erba Mannheim ECL 105 coagulation analyzer, India at KTRH Hematology laboratory. Thawing for subsequent coagulation factor analysis and serial testing of stored fresh frozen plasma and cryoprecipitate was done using Stericox Plasma Thawing Bath, an equipment designed for rapid and uniform thawing of fresh frozen plasma (FFP) bags at 37°C, for 45 mins before the samples are analyzed by Erba Mannheim ECL 105 coagulation analyzer, India and results recorded to assess the coagulation factors changes and levels in cryoprecipitate. Standard storage conditions for the aliquots were observed and maintained to ensure their coagulation factor levels homogeneity.

### Data management and statistical analysis

The data was recorded as numbers (value measured). Statistical analysis was descriptive statistics. The raw data collected was entered in Microsoft office Excel spreadsheet before being transferred to SPSS software version 25.0. The findings were presented in tables and graphs.

### Ethical considerations

Institutional ethical clearance was obtained from Baraton ethical review committee (UEAB/ISERC/02/05/2022) and research permit obtained from National Commission for Science and Technology (NACOSTI) - NACOSTI/P/22/17542.

## Results

The study involved 108 participants who included both male and female. From the analysis, most of the study participants 56 (51.85%) were male blood donors and 52 (48.15%) of blood donors being female. The analysis implies that, majority of the respondents in the study were dominantly male as compared to female as illustrated in the figure below.

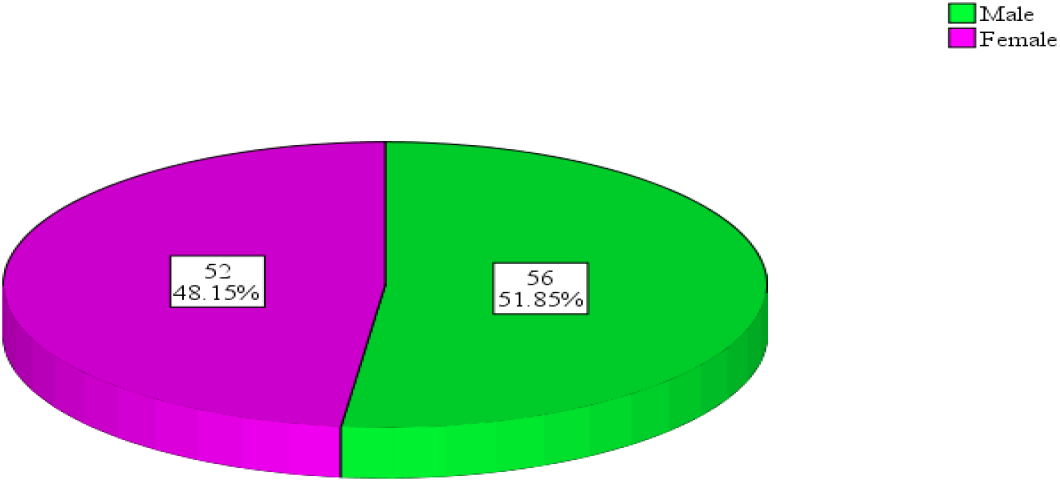

For the realization of the results, Friedman test analysis was employed to establish the changes in coagulation factors in cryoprecipitate plasma. By the use of the mean rank test of the Friedman test analysis, the study was able to establish the differences in time period for coagulation factors in cryoprecipitate plasma of the donated blood. The results were as shown in the table below.

**Table 1:** Friedman’s Test for Mean Rank.

| Variables | Mean Rank |
| --- | --- |
| CRYOPW1 | 3.00 |
| CRYOPW3 | 1.99 |
| CRYOPW5 | 1.01 |

The mean rank for CRYOPW1 to CRYOPW5 was in a decreasing trend with 3.00, 1.99 and 1.01 respectively. This confirms a steady significant difference in mean ranks for the time period though in a decreasing trend thus, the coagulations factors reduce as the time goes by.

The data was further analyzed to establish the changes on coagulation factors in cryoprecipitate to evaluate gender influence on the changes and the results were as shown below;

**Table 2:**
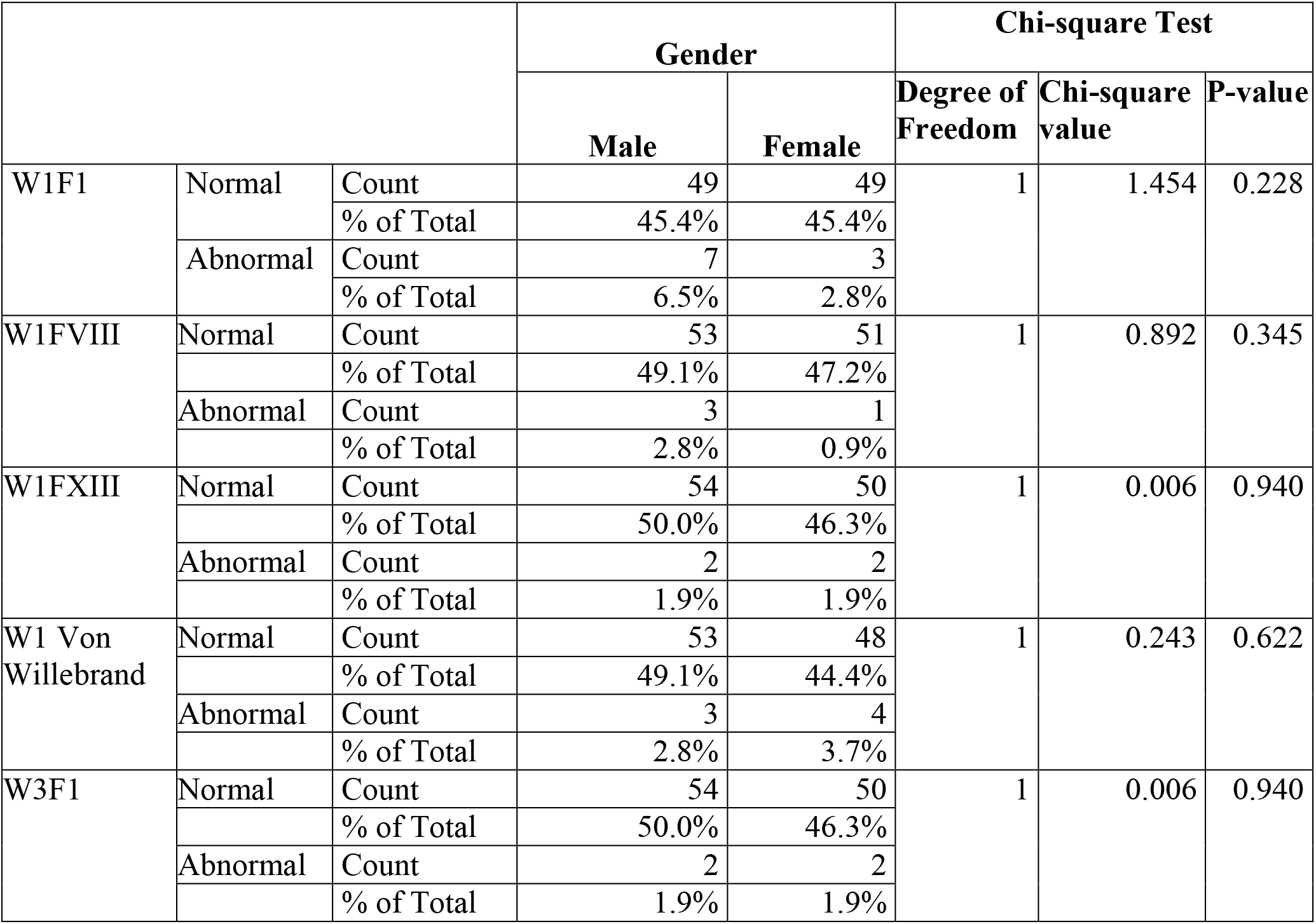
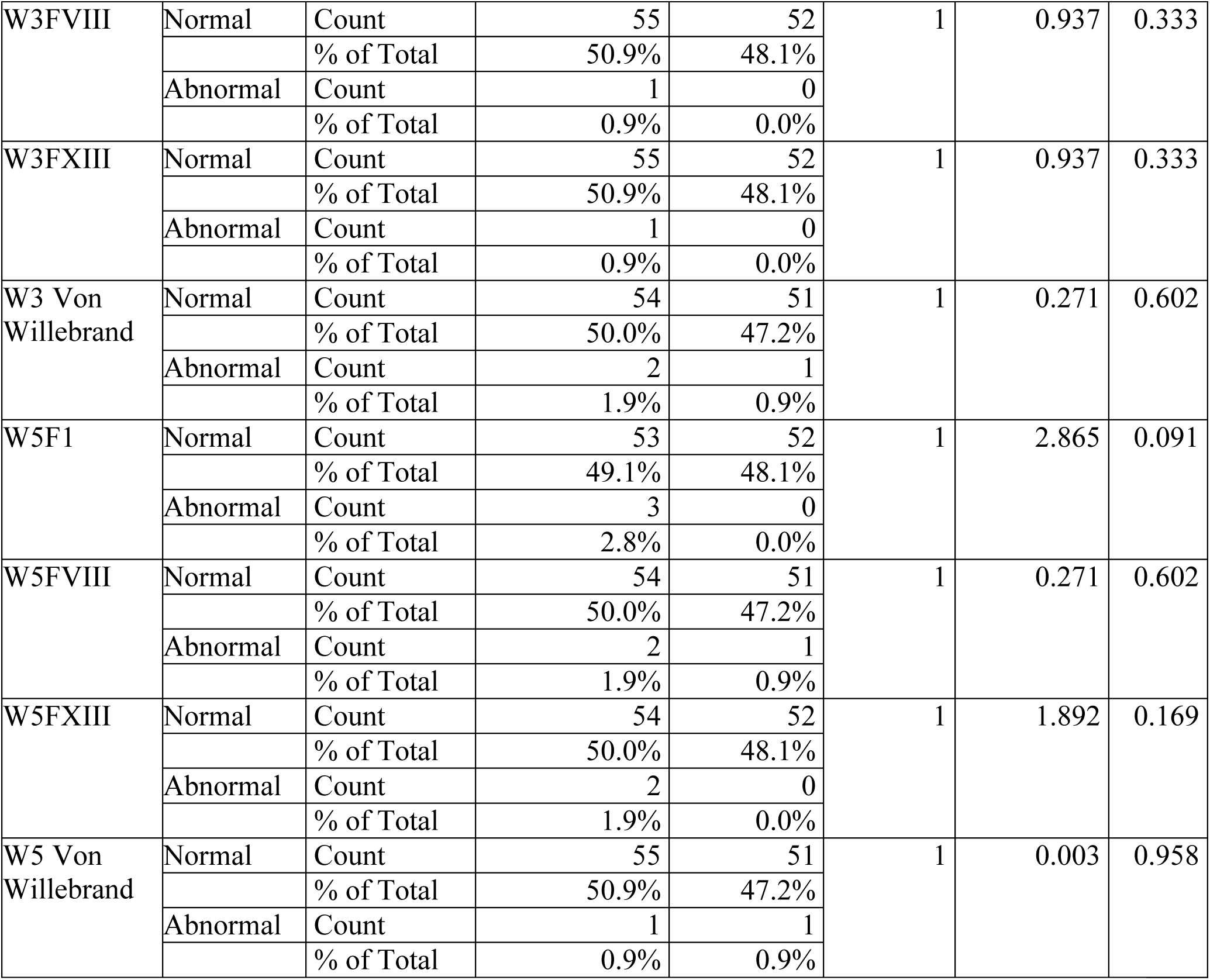
Chi-square Test for Influence of Gender on Coagulation Factors in Cryoprecipitate Plasma.

From the results above, week one Factor one showed that, 49 (45.4%) of male had normal range test while only 7 (6.5%) were abnormal range and 49 (45.4%) representing those female donors who had normal range test with only 3 (2.8%) for the abnormal range test. Week five of the study showed that, 55 (50.9%) were male who had normal range test for the cryoprecipitate plasma with only 1 (0.9%) representing abnormal range and 51 (47.2%) for the female had normal range test with only 1 (0.9%) indicating abnormal range. The coagulation factors in cryoprecipitate plasma in the donated blood were not affected by the gender of the blood donor. This was shown by the results of the p-value as from week one to week five of the study (p-value<0.05). Thus, the gender of the donor has no effect on the coagulation factors in cryoprecipitate plasma.

## Discussion

The mean of coagulation factors in cryoprecipitate plasma for the first week of the study being at 119.10 with a standard deviation of 19.96 with the other weeks recording a change which was significant. This means that most of the coagulation factors in cryoprecipitate plasma across the five-week study did not vary widely, but were distributed around the mean value hence a significant difference in coagulation factors for the period of the study. This finding is anchored by (6) that storing of blood after donation at given storage conditions, the coagulation factors in cryoprecipitate plasma decreases though significantly with a decreasing rate of less ten percent averagely which in support of the study results as shown. The coagulation factors in cryoprecipitate plasma in the donated blood were not affected by the gender of the blood donor. This was shown by the results of the p-value as from week one to week five of the study (p-value>0.05).

## Conclusion

The gender of the donor has no effect on the coagulation factors in cryoprecipitate plasma

## Data Availability

All data produced in the present work are contained in the manuscript

## Data availability

All data are shared on this manuscript

## Acknowledgements

Special appreciation to my mentors and supervisors of this research project, Dr. Benson Nyanchongi (PhD) and Dr. Rodgers Norman Demba (PhD)

Special thanks to lecturers in the department of biomedical science (Kisii University) for the skills and knowledge gained.

Special thanks to KTRH, Kisii University and NACOSTI for the opportunity they gave me to carry out this study.

My special thanks also go to my family, parents (Hellen Ogolla and Joseph Ogolla) and siblings without whom this incredible experience would not have been possible.

